# Pronounced state-level variation in prescription cannabinoids to Medicaid patients

**DOI:** 10.1101/2022.06.04.22275992

**Authors:** Edward Y. Liu, Kenneth L. McCall, Brian J. Piper

## Abstract

Dronabinol is approved for chemotherapy induced nausea as well as vomiting and HIV-induced anorexia, while cannabidiol is primarily approved for childhood epileptic disorders Lennox-Gastaut and Dravet syndrome. The use pattern for these prescription cannabinoids in the US is unknown. This study analyzed Medicaid claims for dronabinol and cannabidiol prescriptions from 2016-2020. Dronabinol prescriptions decreased by 25.3% from 2016 to 2020 while cannabidiol prescriptions increased by 16,272.99 % from 2018 to 2020. The spending on these drugs parallels that on their prescription trend with a 66.3% decrease in reimbursement for dronabinol ($5.7 million in 2020), whereas that of cannabidiol increased by +26,582.0% ($233.3 million in 2020). Dronabinol prescriptions in Connecticut were 136.4 times larger than New Mexico and 17 states had zero prescriptions. Idaho prescriptions, when corrected for number of enrollees, were 15.4-fold higher than Washington DC. Prescribing of dronabinol and cannabidiol did not differ significantly based on the whether the state had an active medical marijuana program. The prescriptions of pharmaceutical grade tetrahydrocannabinol were decreasing while cannabidiol was increasing. This study also identified pronounced state level variation in cannabinoid prescribing to Medicaid patients. Further research to identify the origins of these disparities is needed.

## Introduction

Dronabinol is synthetic tetrahydrocannabinol (THC) that has been US Food and Drug Administration (FDA) approved since 1985 to treat HIV/AIDs-induced anorexia and to patients who do not respond to conventional anti-emetics from the nausea and vomiting of chemotherapy (Badowski and Yanful 2018; May and Glode 2016). Dronabinol activates the CB_1_ receptor, stimulating appetite and provoking an anti-emetic response by reducing the emetic effects from endogenous chemicals like dopamine and serotonin (Badowski and Yanful 2018; May and Glode 2016). Over half of chemotherapy cycles adversely affected the quality of life for patients by inducing undesirable effects including nausea, vomiting, anorexia, nutrient depletion, and metabolic imbalances. By using dronabinol alone or in combination with conventional anti-emetics like ondansetron and prochlorperazine, randomized clinical trials concluded that dronabinol helped prevent nausea and vomiting episodes (May & Glode, 2016). Prescription THC combats weight loss and wasting syndrome from HIV by increasing appetite from the CB_1_ effects (Badowski and Yanful, 2018). The causes of weight abnormalities are hypothesized to be due to several factors, including reduced caloric intake from depression, increased financial difficulties and opportunistic infections, and aggravated metabolic dysfunctions. Dronabinol helped HIV-positive patients increase their daily caloric intake by consuming more meals throughout the day (Haney, Gunderson, Rabkin, Hart et al. 2007).

A randomized double-blind study demonstrated that dronabinol can also treat neuropathic pain associated with multiple sclerosis (Schimrigk, Marziniak, Neubauer, Kugler et al. 2017). Although there were adverse reactions, including restlessness, irritability, sleep interference, decreased appetite, and drug-dependence, this cannabinoid showed promising long-term therapeutic effects in treating neuropathic pain with little to no signs of abuse or dependence. Dronabinol was switched from Schedule II to Schedule III in 1998 with low-moderate potential for abuse due to its gradual onset of action, dysphoria, and other factors (Institute of Medicine 1999). Considerations regarding drug-drug interactions, contraindications, use in specific populations, and other unfavorable effects should be carefully considered. The US FDA (2017) indicated that dronabinol could affect the metabolism of other drugs by inducing or inhibiting cytochrome P450 enzymes (3A4 and 2C9), aggravating mental and/or physical symptoms in patients with neuropsychiatric disorders, inducing fetal harm in pregnant women, causing lactating issues, and exacerbating the neuropsychiatric problems of elderly patients because they are more sensitive to the medications.

The prescription grade of cannabidiol (Epidiolex) has gained recent popularity since its approval by the FDA on June 25, 2018, as the first plant-derived, purified pharmaceutical-grade cannabidiol (CBD) in the US to treat patients ≥ 1 with Dravet Syndrome (DS) or Lennox-Gastaut Syndrome (LGS) which are types of childhood medically refractory epileptic disorders (Abu-Sawwa, Scutt, and Park 2020; Cigna 2022). This drug was initially placed in Schedule V and in April 2020, it was descheduled entirely (DEA Deschedules Antiepileptic, 2020, United States Drug Enforcement Administration 2018). Four randomized, double-blind, placebo-controlled clinical trials were done at 58 sites in Europe and the US to address the efficacy and safety of cannabidiol in treating LGS and DS with 550 patients aged 2-55 (US Food and Drug Administration, 2018a). The GWPCARE1 trial showed a statistically significant decrease in the number of seizures in individuals with DS with an improved quality of life, while the GWPCARE 3 and 4 trials demonstrated a statistically significant dose-dependent effect on reducing the number of seizures in individuals with LGS, with approximately half of patients demonstrating ≥50% reduction in seizures (Abu-Sawwa, Scutt, and Park 2020). The study concluded that cannabidiol should be used as an adjunct with other anti-epileptic medications to combat DS and LBS. The drug-drug interactions however must be carefully considered, as cannabidiol inhibits CYP2C8, CYP2C9, CYP2C19, UGT1A9, and UGT2B7 and there are dose-dependent increases in adverse effects from the adjunct therapy including lethargy, somnolence, fatigue, nausea, vomiting, and increased liver enzymes and frequency of upper respiratory infections (US FDA, 2018b).

The US FDA (2020) specified that the efficacy and safety use of prescription cannabidiol has led to its approval in 2020 to treat another epileptic disease called tuberous sclerosis, a rare genetic disorder that causes benign tumors to grow in the brain and other parts of the body, resulting in various symptoms including seizures, developmental delays, and other abnormalities. Although the benefits and the safety of the drug are promising as indicated by the results of the multicenter clinical trials, further investigation is still needed before unqualified endorsements for long-term usage are made due to limited data in the drug-drug interactions, its reproductive adverse health effects, and other problematic side effects (Abu-Sawwa, Scutt, and Park 2020). The mechanism in which cannabidiol induces anti-seizure effects remains unknown, but is theorized to alter neurochemicals, such as serotonin, gamma-amino butyric acid, t-type calcium channels, and N-methyl-D-aspartate by binding to CB_1_ and CB_2_ receptors. CB_1_ is most dense in the basal ganglia, cerebellum, cortex, and hippocampus, which contributes to diseases affecting neurological conditions related to altered brain reward mechanisms, processes of learning and memory, as well as mood and anxiety disorders, whereas CB_2_ is present primarily in immune and hematopoietic systems, lacking psychoactive effects (Pacher, Bátkai, and Kunos 2006). The cannabidiol may interact with several signaling systems incluing the Transient Receptor Potential Vanilloid type 1 (TRPV-1) to induce anticonvulsant effects (Silva, Del Guerra, Lelis, and Pinto 2020).

Since their approval, dronabinol and cannabidiol have also been examined for various off label uses (Abu-Sawwa, Scutt, and Park 2020; Badowski and Yanful 2018; National Academies of Sciences, Engineering, and Medicine 2017). However, there is currently a lack of information about their use patterns in the US. This study analyzed the prescriptions for dronabinol and cannabidiol throughout the US from 2016 to 2020 for Medicaid patients and quantified their state-level variation.

## Materials and Methods

### Procedures

The national and state-level number of prescriptions for cannabidiol (Epidiolex) and dronabinol (Marinol, Syndros) were obtained from Medicaid (Centers for Medicare and Medicaid Services 2022a; IBM Watson Health 2022) for 2016 to 2020. Medicaid is a federal and state insurance program that provided healthcare coverage in 2020 for 82 million low-income patients (Centers for Medicare and Medicaid Services 2022b). This year was chosen as the end period for analysis because it was the last year for which full drug data was available when analysis was completed (3/2022). The mean number of prescriptions for each cannabinioid from 2016 to 2020 was determined. A comparison was made for cannabinoid prescribing based on whether the state had a medical marijuana law and dispensaries active selling cannabis in 2020 (Marijuana Policy Project 2022). Finally, the total drug reimbursement was assessed.

### Data Analysis

The analyses were: (1) Heat maps among the 50 states and Washington DC for both cannabinoids (Babicki, Arnd, Marcu, Liang et al. 2016); (2) total ratio of highest to lowest (non-zero) prescriptions per state, corrected for the number of Medicaid enrollees in 2020 in the US with states outside a 95% confidence interval (mean ± 1.96 *SD) interpreted as statistically significant (*p* < .05); (3) correlation between the number of dronabinol and cannabidiol prescription among the states; (4) the average total number of prescriptions of both cannabinoids from 2016 to 2020 among all the states and the population corrected prescriptions for both cannabinoids at the state and national levels; (5) a t-test to examine for a plausible association between the number of prescriptions for both cannabinoids in states that had or had not have between states that had, and had not, approved medical marijuana with ongoing dispensary sales, and (6) the annual total drug reimbursement ($) from 2016 to 2020 for both cannabinoids. The data analyses and figures were completed using GraphPad Prism v 9.3.1.

## Results

Dronabinol average prescriptions per state showed a gradual decrease (−25.3%) from 2016 to 2020 (Supplemental Figure 1A). Similarly, when prescriptions were corrected for the number of enrollees, there was a 31.6% decrease (Supplemental Figure 1B). There was no difference in dronabinol prescribing in 2020 between states that had, and had not, approved medical marijuana (Supplemental Figure 2). Total spending on dronabinol decreased 66.3% from $17,058,455.46 in 2016 to $5,748,587.88 in 2020 (Supplemental Figure 3).

Dronabinol prescriptions in the top state, Connecticut (180.0), was 136.4-fold greater than that of the lowest non-zero state (New Mexico = 1.3, Figure 1). Connecticut, Nebraska, Missouri, and Maine had a greater number of prescriptions (> 1.5 SDs) than the national mean. One-third of states (17) had no dronabinol prescriptions including a cluster (NV, ID, MT, WY, ND, SD, IO, WI) in the north-central US.

**Figure 1.**
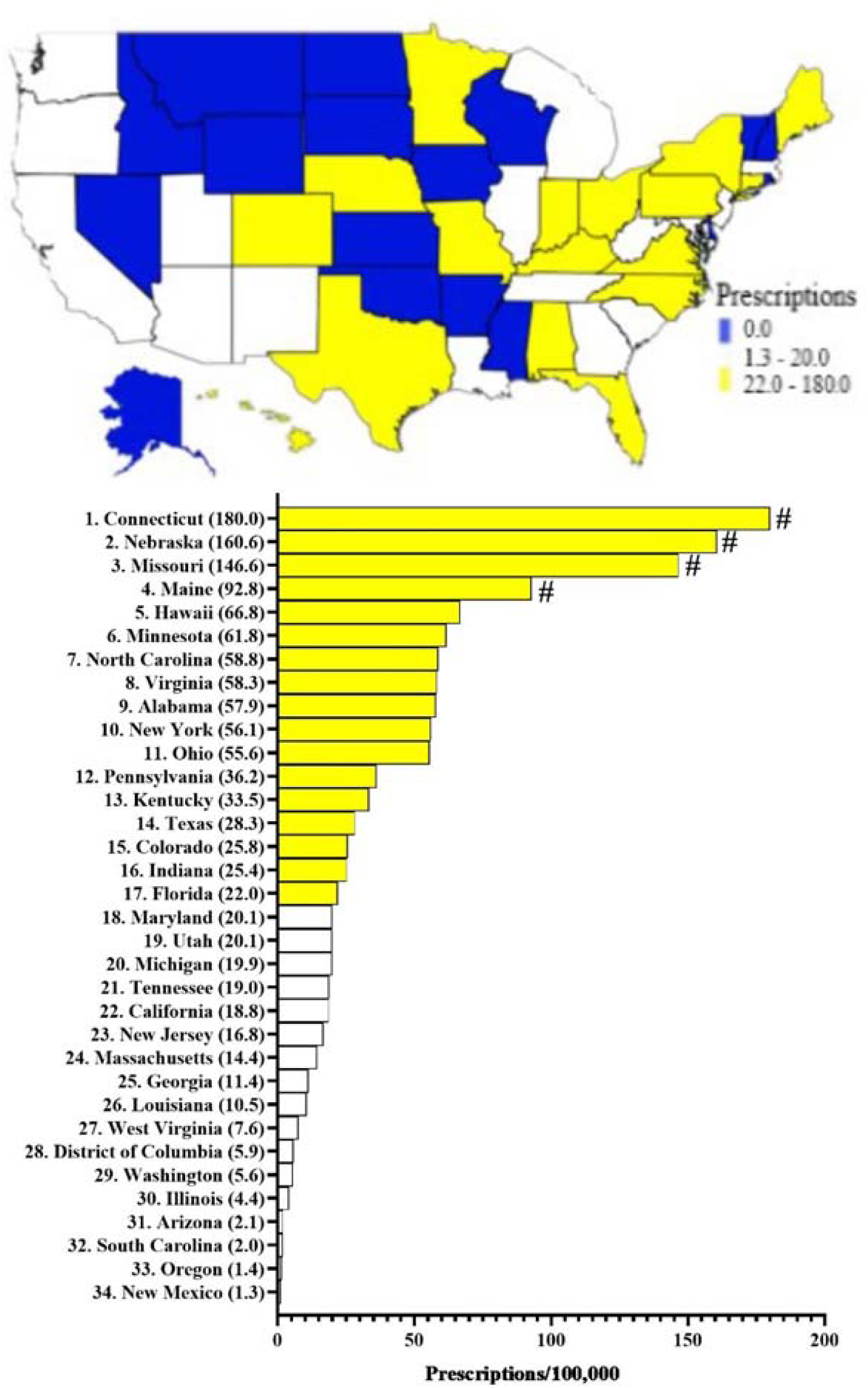
Dronabinol prescriptions per 100,000 Medicaid enrollees. Heatmap (top) and ranked (bottom) in 2020. States outside ^#^1.50 Standard Deviations from the mean. Seventeen states including DC with 0 prescriptions are not shown on the bar graph.

Cannabidiol average prescriptions showed a pronounced (+16,272.99%) increase since its approval in 2018 until 2020 (Supplemental Figure 1C). Similarly, the number of prescriptions when corrected for the number of enrollees showed a drastic increase (+14,496.77%) during this interval (Supplemental Figure 1D). Medicaid spending also demonstrated an exponential increase of 26,581.98% from $874,370.82 in 2018 to $233,299,416.18 in 2020 (Supplemental Figure 2).

Figure 2 shows a 15.4-fold difference in population corrected prescription of cannabidiol between Idaho and DC in 2020. Idaho, Wyoming, North Carolina, and New Hampshire demonstrated significantly greater prescribing than the average. Florida showed greater (> 1.5 SD) while Alaska, New Mexico, Hawaii, and DC had lower (< 1.5 SD) prescribing than the average. However, there was no difference in cannabidiol prescribing in 2020 between states that had, and had not, approved medical marijuana (Supplemental Figure 2)

**Figure 2.**
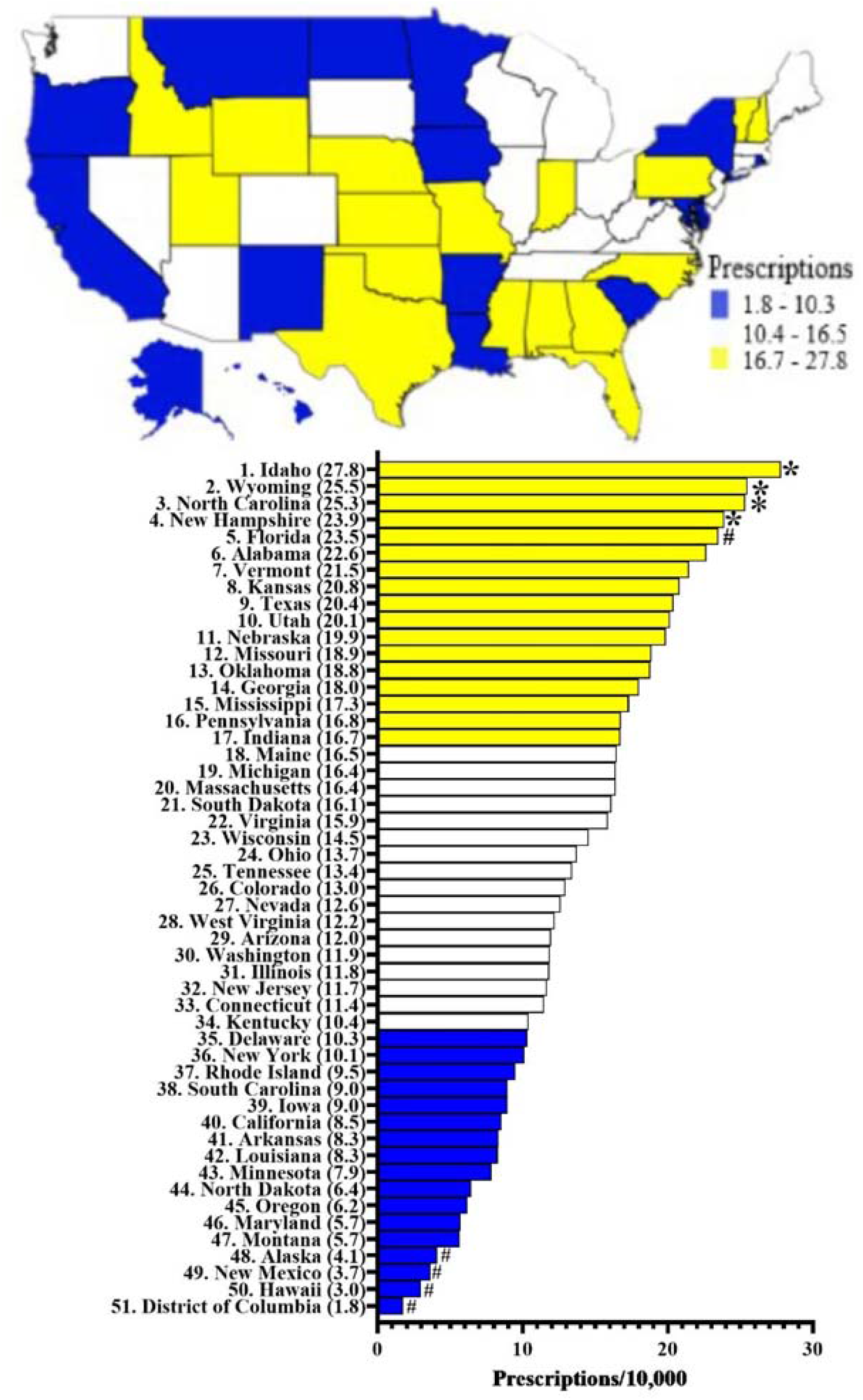
Cannabidiol prescriptions per 10,000 Medicaid enrollees. Heatmap (top) and ranked (bottom) in 2020. (States outside *1.96 or ^#^1.50 Standard Deviations from the mean).

## Discussion

There was a 25% decrease in dronabinol prescriptions between 2016 and 2020 and an exponential increase in cannabidiol prescriptions of 16,273% from 2018 to 2020, with pronounced state variation in prescriptions to Medicaid patients. Dronabinol can be used alone or in combination with other drugs to treat chemotherapy-induced nausea and vomiting, HIV-associated anorexia, and chronic pain since its approval in 1985 (O’Donnell 2021), whereas cannabidiol has specifically been proven to work in adjunct with other antiepileptics to treat rare epileptic disorders, specifically Lennox Gaustaut and Dravet Syndromes since its approval in 2018 (Abu-Sawwa, Scutt, and Park 2020). Understanding the distribution of the two medical cannabis provides a novel insight into their pharmacoepidemiology, especially with the rising attention in the use of cannabis to combat various medical conditions (National Academy of Sciences, 2017).

The sizeable state-level variation in dronabinol prescriptions may be explained by various factors including differing Medicaid Preferred Drug List (PDL) policies (Kaiser Family Foundation 2019). However, the presence of an active medical marijuana program in a state did not impact dronabinol prescribing (Mahabir, Smith, Vannabouathong, Merchant et al. 2021; Marijuana Policy Project 2021). Additionally, the lower number of dronabinol prescriptions compared to cannabidiol might be because dronabinol is a Schedule III drug, is less likely to be indicated as a preferred drug in the Medicaid PDL or included in it, and the cannabis has step therapy policies in some states (Agency For Health Care Administration 2022; Magellan Health 2020; Texas Health and Human Services 2022; United States Drug Enforcement Administration 2018). PDLs are generally implemented by states to reduce prescription costs, but the application of this policy varies among the states depending on factors such as the possible effects of drug restrictions on the quality of care, issues between cost-savings and quality of care, and the administrative costs related to the program (Gifford, Winter, Wiant, Dolan et al. 2020; Ovsag, Hydery, and Mousa 2008). According to Ovsag, Hydery, and Mousa (2008), the inclusion of certain drugs into the PDL is specifically determined by the negotiation of the price rebates between the manufacturer and the state Medicaid programs. Step-through parameters like the required use of 5-HT_3_ receptor antagonists for cancer-associated nausea and vomiting prior to approval of dronabinol might explain the larger variability of dronabinol prescriptions among the states (Magellan Health 2022).

Similarly, the pronounced increase in cannabidiol prescriptions from 2018-2020 may be explained increased incorporation into the PDL of some states (“Dea Deschedules Antiepileptic” 2020; Magellan Health 2020; Texas Health and Human Services 2022) and its de-scheduling in 2020. The high population corrected cannabidiol prescriptions in Idaho and Wyoming might be explained by the late or lack of Medicaid expansion, especially since states that have expanded Medicaid enable individuals who are under 65 to qualify for the program if their income is up to 138% of the federal poverty level (Kaiser Family Foundation 2022). Idaho expanded its Medicaid program in 2020, whereas Wyoming has not (Healthinsurance.org 2022). A previous study has shown that the prevalence of epilepsy is higher in low-income Americans, individuals with pre-existing disabilities, and comorbid conditions than that of the general population (Kaiboriboon, Bakaki, Lhatoo, and Koroukian 2013). Therefore, states that were late to expand or still have not expanded may have increased use of cannabidiol, specifically in populations with poorer health and lower income than states who have expanded Medicaid.

In addition to the differing policies, regulations, and medical marijuana legalization status among the states, the varied knowledge of the use, benefits, and adverse effects of medical marijuana combined with the negative stigma of the substance and inadequat scientific evidence on the medication’s safety and efficacy may also contribute to the varying amounts of prescriptions of dronabinol and cannabidiol (Ronne, Rosenbaek, Pedersen, Waldorff et al. 2021).

The drug reimbursement for Medicaid parallels that of the temporal trend in prescriptions for both cannabinoids. The Red Book from IBM Micromedex (2022) are generally used as price benchmarks for drugs, and it lists the average wholesale price (AWP) and wholesale acquisition cost (WAC) of dronabinol as $4,242 and $1,213.8, respectively. The AWP and WAC are $1,848.00 and $1,540.00, respectively for a 100 mL oral solution of Epidiolex. The annual price of cannabidiol ($32,500) is non-trivial although this is aligned with the cost of other anti-epileptics (Kheen and El-Hajj, 2022). The sales of non-pharmaceutical grade cannabidiol in the US rose from $535 million in 2018 to $4.6 billion in 2020, and the growth is expected to rise to more than $20 billion by 2024-2025 (Sill 2021; Statista 2021). This greatly dwarfs the $233.3 million spent by the Medicaid program in 2020 for pharmaceutical grade cannabidiol.

Some limitations and future directions are noteworthy. These novel findings are limited to the 82 million patients served by the Medicaid program. Future research of Medicare or privately insured US patients or in other countries is warranted. The Medicaid State Drug Utilization database provides information about prescribing but further study with electronic medical records would be necessary to further characterize prescribing patterns. As the approved indications for cannabidiol (DS incidence 1 per 22,000-40,900, LGS was 4% of all cases of childhood epilepsy, Tuberous Sclerosis incidence ranges from 1 per 5,800 to 10,000 live births) are relatively rare, further research to characterize the off-label use of cannabidiol may be informative.

## Conclusion

The number of dronabinol prescriptions showed a moderate decrease, while cannabidiol demonstrated an exponential increase since its 2018 approval. There was also pronounced state-level variation in prescriptions of these cannabinoids. Further research needs to be completed to determine which factors including PDL account for these regional differences.

## Data Availability

All data produced are available online at: https://www.medicaid.gov/medicaid/prescription-drugs/state-drug-utilization-data/index.html

## Acknowledgments

We thank Raymond Stemrich, MHA for technical assistance. Software used in this study was provided by National Institute of Environmental Health Sciences (T32 ES007060-31A1). BJP was supported by the Health Resources Services Administration (D34HP31025).

## Declaration of Interest Statement

BJP was part of an osteoarthritis research team (2019-2021) supported by Pfizer and Eli Lilly. The other authors have no disclosures.

---

**Supplemental Figure 1.**
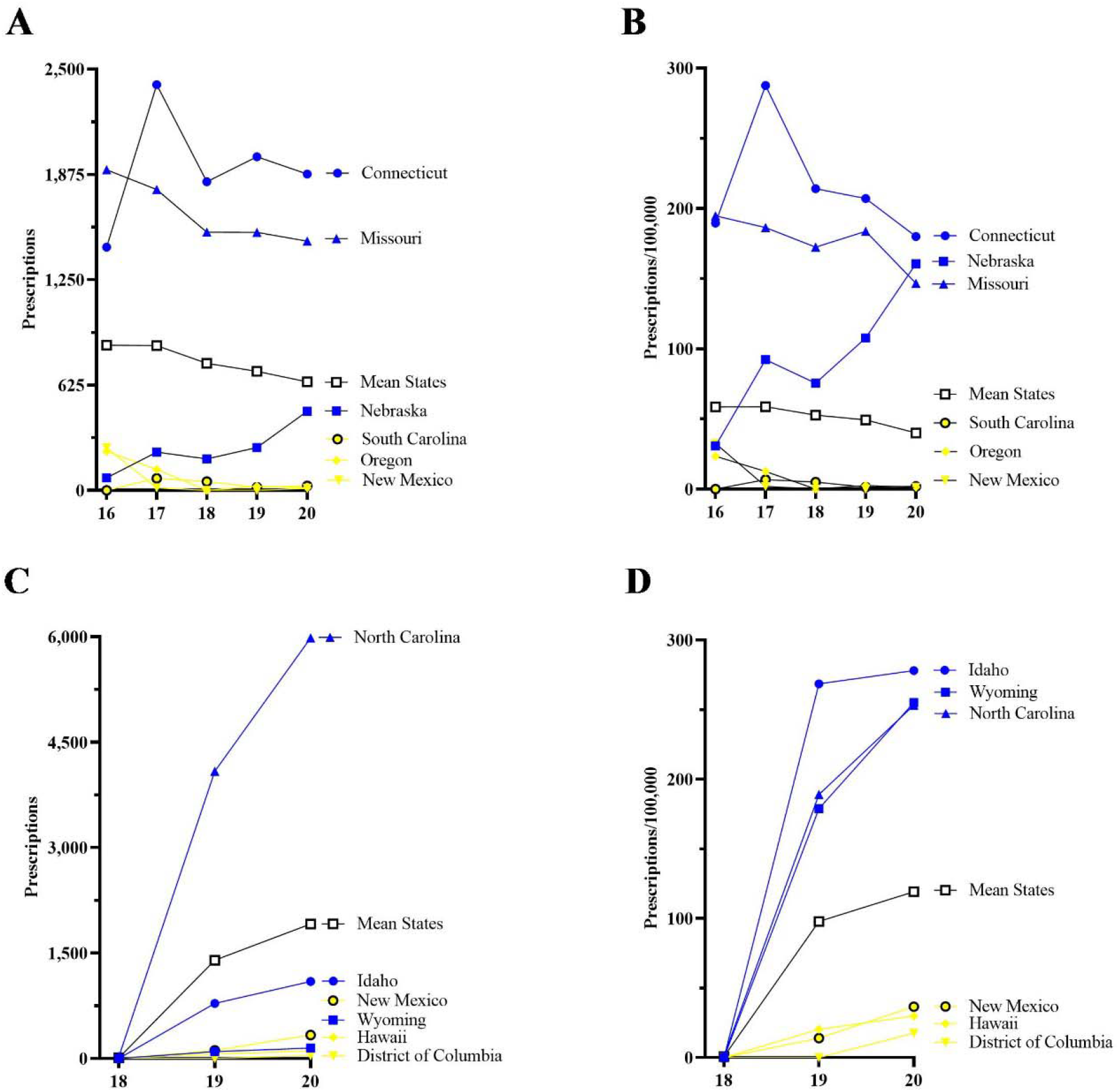
Total number of prescriptions (A, C) and corrected for number of enrollees (B, D) for dronabinol (A, B) and cannabidiol (C, D) to Medicaid patients for the three highest and three lowest states for 2016 to 2020. The mean number of prescriptions among 50 states and Washington DC are also shown.

**Supplemental Figure 2.**
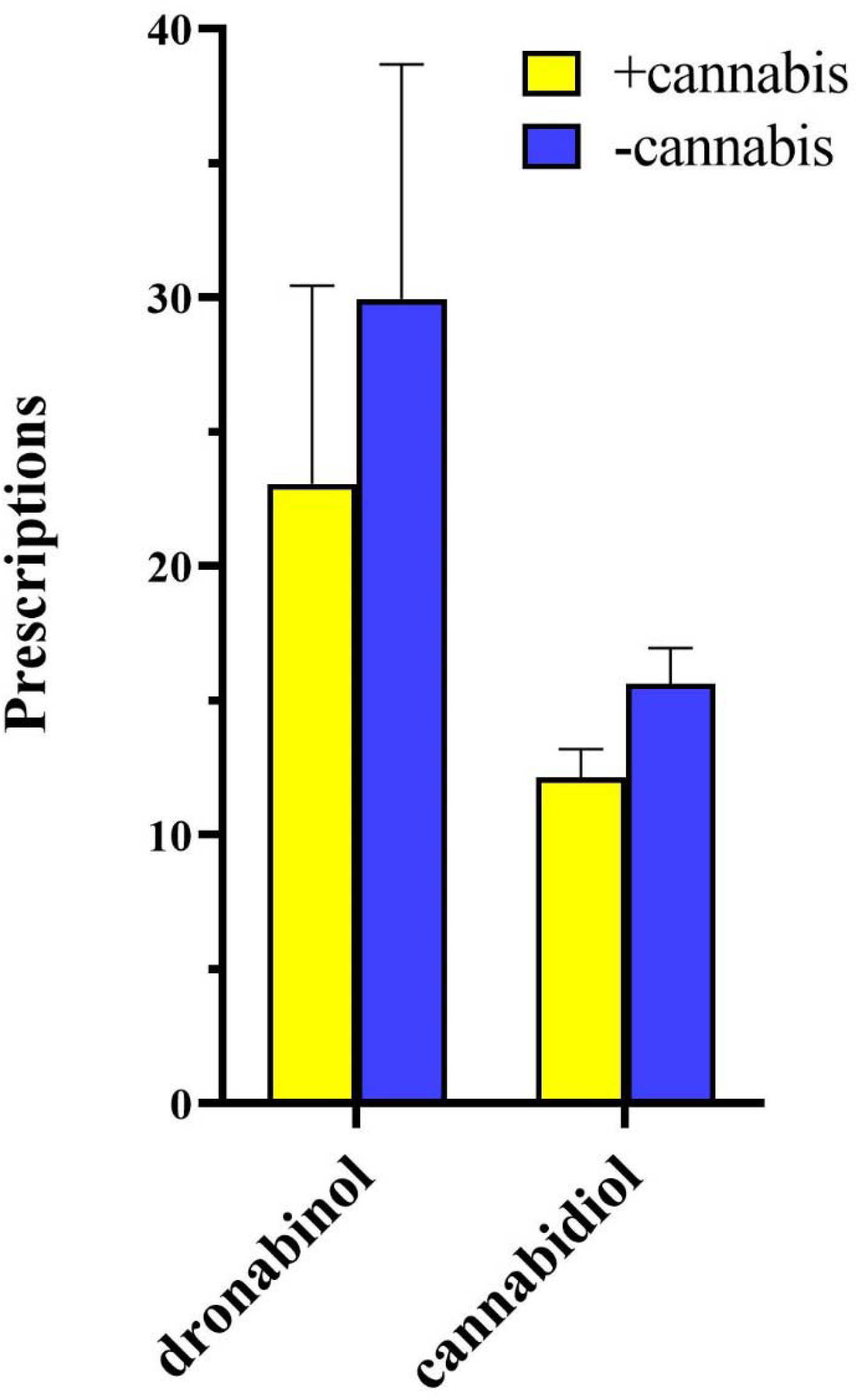
Comparison between the population corrected average prescriptions of dronabinol and cannabidiol in states that do (+) and do not (-) have medical cannabis in 2020.

**Supplemental Figure 3.**
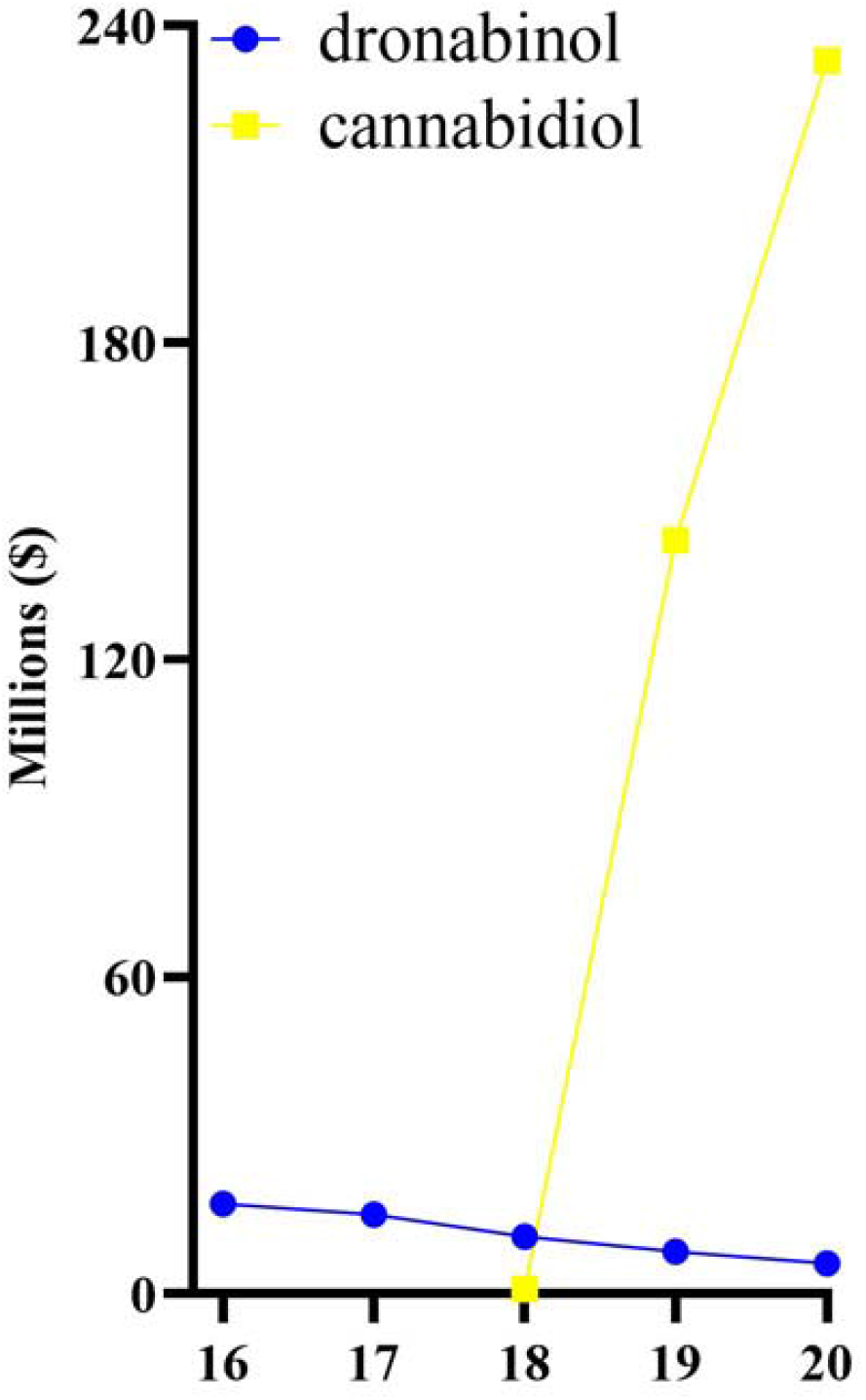
The total drug reimbursement in Medicaid of dronabinol and cannabidiol in millions ($) from 2016 to 2020.

## Notes

### Author Declarations

The IRBs of the University of New England and Geisinger approved this research as exempt.

